# Quantifying Epistemic and Aleatoric Uncertainty in 3D U-Net Segmentation

**DOI:** 10.1101/2021.09.20.21263844

**Authors:** Craig K Jones, Guoqing Wang, Vivek Yedavalli, Haris Sair

## Abstract

This work shows a derivation of a multinomial probability function and quantitative measures of the data and epistemic uncertainty as direct output of a 3D U-Net segmentation network. A set of T1 brain MRI images were downloaded from the Connectome Project and segmented using FMRIB’s FAST algorithm to be used as ground truth. A 3D U-Net neural network was trained with sample sizes of 200, 500, and 898 T1 brain images using a loss function defined as the negative logarithm of the likelihood based on a derivation of the definition of the multinomial probability function. From this definition, the epistemic (model) and aleatoric (data) uncertainty equations were derived and used to quantify maps of the uncertainty in data prediction. The epistemic and aleatoric uncertainty decreased based on the increasing number of training data used to train the neural network. The neural network trained with 898 volumes resulted in uncertainty maps that were high primarily in the tissue boundary regions. The uncertainty was averaged over all test data (connectome and tumor separately) and the epistemic uncertainty showed a decreasing trend, as expected, with increasing numbers of data used to train the model. The aleatoric uncertainty showed a similar trend, but it was less obvious, which was also expected as the aleatoric uncertainty is not expected to be as dependent on the number of training data. The derived data and epistemic uncertainty equations from a multinomial probability distribution are applicable for all 2D and 3D neural networks.

## Introduction

Neural network segmentation has fast become one of the important uses for machine learning in medical imaging. Studies have used networks to segment brain tumors (Chen et al. 2020; Hussain, Anwar, and Majid 2018; Badža and Barjaktarović 2020; Reddick et al. 1998; Sharif et al. 2020; Dand¹l and Karaca 2021), tumors of the head and neck (Fu et al. 2020; Gou et al. 2020; Park et al. 2019), organs in the abdomen (Edwards et al. 2020; Jiang et al. 2017; Selver 2014; Rafiee, Masoumi, and Roosta 2009; Lee, Chung, and Tsai 2003; Hu et al. 2017). The most popular neural network model for segmentation is the U-Net model (Ronneberger, Fischer, and Brox 2015) and its variants, including U-Net++ (Z. Zhou et al. 2018; Milletari, Navab, and Ahmadi 2016), V-Net (Lin et al. 2019), 3D U-Net (Çiçek et al. 2016) and 3D U^2^-Net (Huang et al. 2019). The U-Net family of segmentation models takes in an image or volume and uses a set of steps to spatially down-sample the input and encode in an increasing number of channels, followed by an up-sampling scheme with the same number of steps to the full spatial resolution. The output of the network typically is a mask of the segmented region.

Deep learning must be accurate and stable to remain reliable (Antun et al. 2020) for use in medical image diagnosis. In the Antun paper, they compared image reconstruction techniques and found the results were not stable to changes (e.g., noise or small perturbations in the image) to the input image. For medical image segmentation the errors may arise at the boundary of regions or where large regions are mislabeled. (Pai et al. 2017). Given potentially significant adverse consequences of inaccurate labeling for medical images, an estimate of the reliability of segmentations is needed if such AI tools are to be used in medical decision making(Senge et al. 2014).

There are two types of uncertainty in measuring a parameter(Hüllermeier and Waegeman 2021): (1) aleatoric uncertainty refers to the variability of an outcome due to inherent random effects and cannot be reduced based on more data; (2) epistemic uncertainty is caused by lack of knowledge (shows what the model does not understand about the data) and can be decreased by adding more training samples. Each of these types of uncertainty are present in all predicted parameters from models, including trained neural networks(Hüllermeier and Waegeman 2021).

Recently, there have been papers that have shown techniques to provide measures of uncertainty in the results of neural network segmentations. Currently, techniques for providing measures of uncertainty rely on: (1) Bayesian inference with placing prior distributions over hierarchical models (Gelman et al. 2008), (2) Bayesian deep learning which places priors over network weights (Kingma et al. 2015). Previously, Senge(Senge et al. 2014) showed classifiers can distinguish between aleatoric and epistemic uncertainty and more recently Amini (Amini et al. 2019) derived the epistemic and aleatoric uncertainty from the learned evidential distribution and applied it directly to a classification network. The Amini paper showed a classification model was able to produce not just the classification but also the data and epistemic uncertainty from within the model outputs.

Our paper has furthered the quantification of uncertainty from a classification neural network (Amini et al. 2019; Senge et al. 2014) by deriving the epistemic and aleatoric uncertainty for a multi-class segmentation model and applied it to normal appearing MRI datasets and a set of brain tumor data. Aleatoric and epistemic uncertainty maps were created for a test set of segmentation data and for the tumor data. The aleatoric and epistemic uncertainties were averaged for each tissue over all test data to compare the changes over the number of training sets for the model. We hypothesize that aleatoric and epistemic uncertainty maps can be generated on U-net based segmentation of medical imaging, specifically MRIs of the brain.

## Methods

### Data

The neural network segmentation was trained and tested based on T1 MRI images. The training data consisted of 928 T1 MRI images from the 1000 Connectome Project (https://www.nitrc.org/projects/fcon_1000/ from locations AnnArbor, Atlanta, Baltimore, Bangor, Beijing, Berlin, Cambridge, Cleveland, Dallas, ICBM, Leiden, Milwaukee, Munchen, Newark, NewHaven, and NewYork). The downloaded data consisted of skull-stripped data in NIFTI format and were downloaded to a local machine. The T1 data was then segmented into WM, GM and CSF masks using FMRIB’s Automated Segmentation Tool (Zhang, Brady, and Smith 2001) with parameters *fast -S 1 -t 1 -n 3*. The automated segmentation from FAST were reviewed by a board-certified radiologist to confirm accuracy. The 1000 Connectome Project data was used for this project as it is an open source dataset, is very large, and has a consistent T1 acquisition.

A second set of T1 weighted data was used as a second test for the prediction and uncertainty and to visualize the epistemic and aleatoric uncertainty in the tumor region which was not a specific class used in training. This data consisted of a set of 10 T1 weighted scans acquired pre-operatively of patients who had visible brain tumors as part of their standard of care. This data was used retrospectively and was approved by the institute’s IRB. The only eligibility criteria is that there was a visible tumor in the T1 MRI images. The data was acquired on a Siemens 3T MRI scanner with parameters TR/TI/TE/flip angle = 2300 / 900 / 3.5 / 9º. The DICOM data was de-identified using internal software based on the RSNA MIRC Clinical Trials Processor (https://mircwiki.rsna.org/index.php?title=CTP-The_RSNA_Clinical_Trial_Processor) de-identification standard and then converted to NIFTI format using `dcm2niix` (Li et al. 2016). For this dataset, skull stripping and FAST segmentation were not performed.

### Neural Network Training

The 3D segmentation neural network used in this work was a convolutional neural network (CNN) based on a 3D U-Net (Udon n.d.; Çiçek et al. 2016) implementation in GitHub (https://github.com/UdonDa/3D-UNet-PyTorch/blob/master/src/model.py) consisting of five encoding and five decoding steps. The U-Net was initialized with random weights based on the default random algorithm in PyTorch. A set of 30 datasets were held as a consistent test (prediction) set leaving 898 datasets for training. The network was trained three times, first with N=200 datasets, second with N=500 datasets and lastly with N=898 datasets. The 500 datasets were chosen randomly from the 898 and the 200 datasets were chosen randomly from the 500. The goal was to minimize results that were dependent on the actual training set. All data randomization was done at the patient level. For each of the training a set of 30 data were randomly chosen for validation. The U-Net training used minimal data augmentation (1% perturbation on the signal intensity), Adam optimizer, learning rate of 0.0001, batch size of 3 and 140 epochs. The loss function is derived from a multinomial distribution of classes and is defined as the negative log-likelihood:

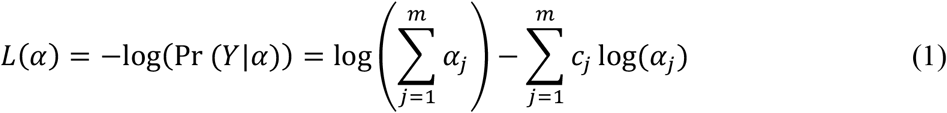

A full derivation of the loss function is shown in the Appendix. After each training, the weights were saved to be used during the prediction method. The Dice coefficient, defined as

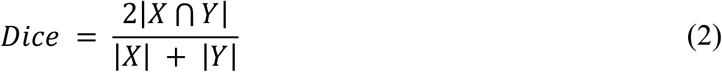

was used as the mask overlap accuracy measure. The implementation was in PyTorch version 1.8 (Paszke et al. 2019) and trained using a Titan XP GPU with 12GB RAM.

### Neural Network Predictions

The Connectome test data held out was passed through each trained neural network and maps of the predicted segmentation, aleatoric uncertainty (Equation 4) and epistemic uncertainty (Equation 5) were created. The model output the values ***α***, was the prediction of being within each tissue. The total *S*_*α*_ was quantified by summing the tissue predictions, from the model, based on Equation 3. The epistemic uncertainty was quantified from Equation 5 for tissue class *j* from the tissue’s predicted probably, *α*_*j*_ and *S*_*α*_. Similarly, the aleatoric uncertainty was quantified using Equation 4 and based on the tissue’s predicted probability *α*_*j*_ and *S*_*α*_. The T1 brain tumor data was passed through each trained neural network and the same prediction and uncertainty maps were created.

### Comparison to Cross Entropy Loss

A copy of the neural network was trained on the same network structure and optimizer but a cross entropy loss function, defined in PyTorch (v1.8), instead of the loss defined in this paper. The network was trained and the final dice scores of the test data were compared to the final dice scores of the test data from the network trained on the loss function defined in this paper.

## Results

### Training Results

The loss and accuracy (Dice) curves were quantified and the set from the N=898 training is shown in Figure 1. The training loss (blue) and validation loss (orange) decayed smoothly over the 140 training epochs (Figure 1, top left). The accuracy curve (mean over the three tissues) increased smoothly up to approximately 0.73 (Figure 1, top right). The training accuracy was plotted for each of the tissues to confirm the tissue accuracy curves were not disparate. The CSF had the lowest training accuracy (Dice score) of 0.64, the WM followed the mean accuracy, and the GM had the best accuracy of 0.83 (Figure 1, bottom left). The validation accuracy as a function of epoch and tissue type had a similar trend to the training accuracy, and is shown in Figure 1, bottom right).

**Figure 1:**
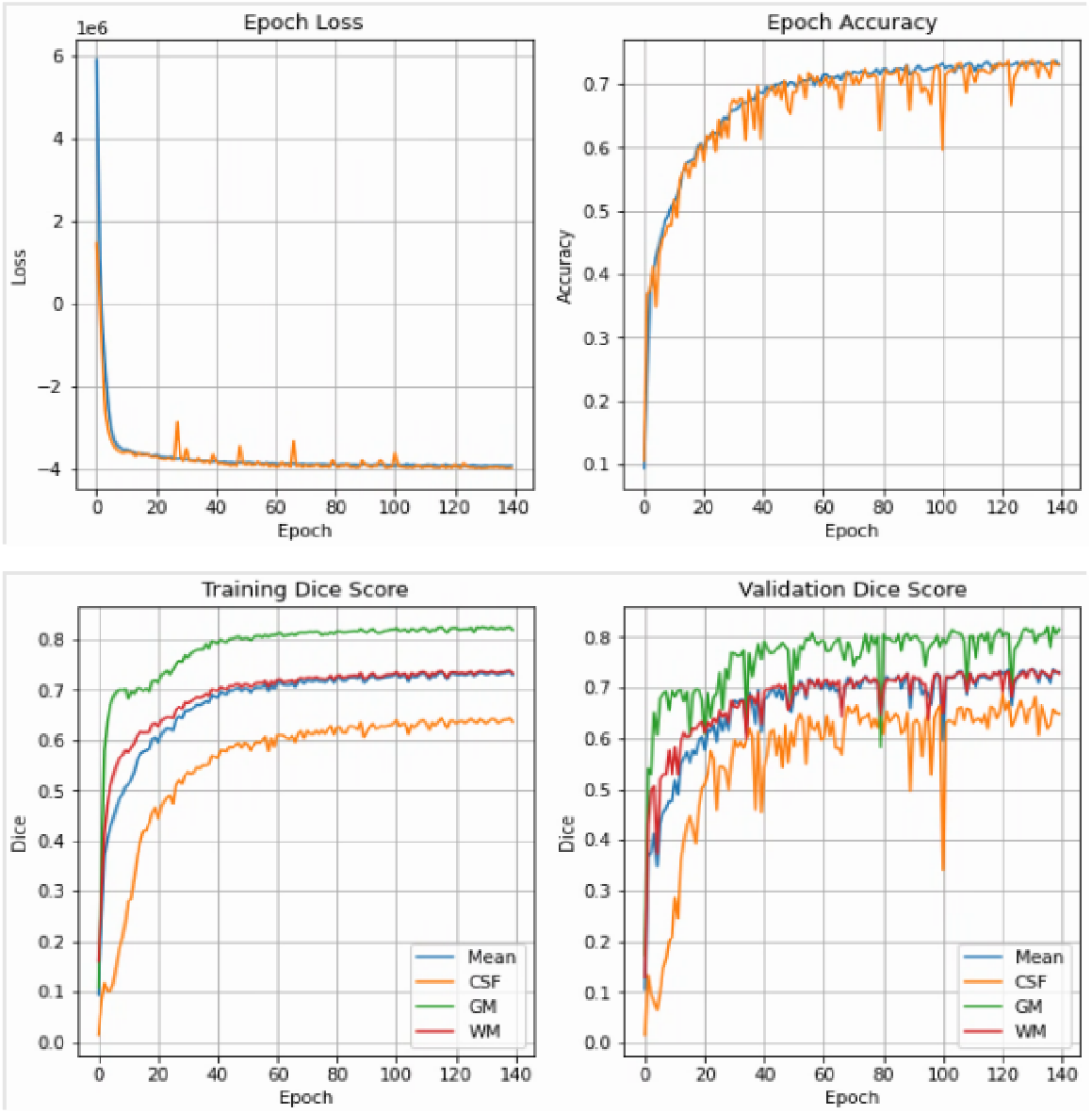
Training loss (top left), accuracy (top right) along with the training dice score per tissue type for the training (bottom left) and validation data (bottom right). The data shown here is for the N=898 training session.

### Connectome Data Results

An example segmentation for one slice of one test dataset is shown in Figure 2. The top row shows the tissue segmentation classes based on predictions from the N=200, N=500 and N=898 trained models, followed by the original T1 image. Generally, the tissue segmentation visually appears more accurate for the N=898 segmentation. The following panels have the aleatoric and epistemic uncertainty for each of the Background, CSF, WM, and GM classes. The colormaps are scaled such that all are between 0 and 0.200 for easy comparison. The aleatoric uncertainty overall had an uncertainty value of approximately 0.075, or less, for all three tissues. Generally, the uncertainty lay at the border of the tissue boundaries. The epistemic uncertainty decreased spatially with more data used to train the network and for the N=898 predictions the highest uncertainty was primarily in the tissue boundaries. Some of the highest uncertainty lay in the regions of the caudate nucleus, internal capsule anterior horn and then putamen. Overall, the epistemic uncertainty decreased as the number of data used to train the model increased. For example, in the white matter uncertainty maps, the larger white matter regions decrease from approximately 0.100 (N=200), to 0.05 (N=500) to near zero (N=898). This is consistent with the idea that the epistemic uncertainty will decrease with an increase in the number of data used for training. The trend is slightly less clear in the GM epistemic uncertainty maps as the GM ribbon around the cortex is thin and therefore will have partial volume with the WM. But, even with it being thin, there is a decrease in the epistemic uncertainty from 0.175 (N=200) to 0.125 (N=500) to near 0.100 (N=898). Along with the quantitative decrease, there is clearly a visual difference in the GM epistemic uncertainty maps with increasing N.

**Figure 2:**
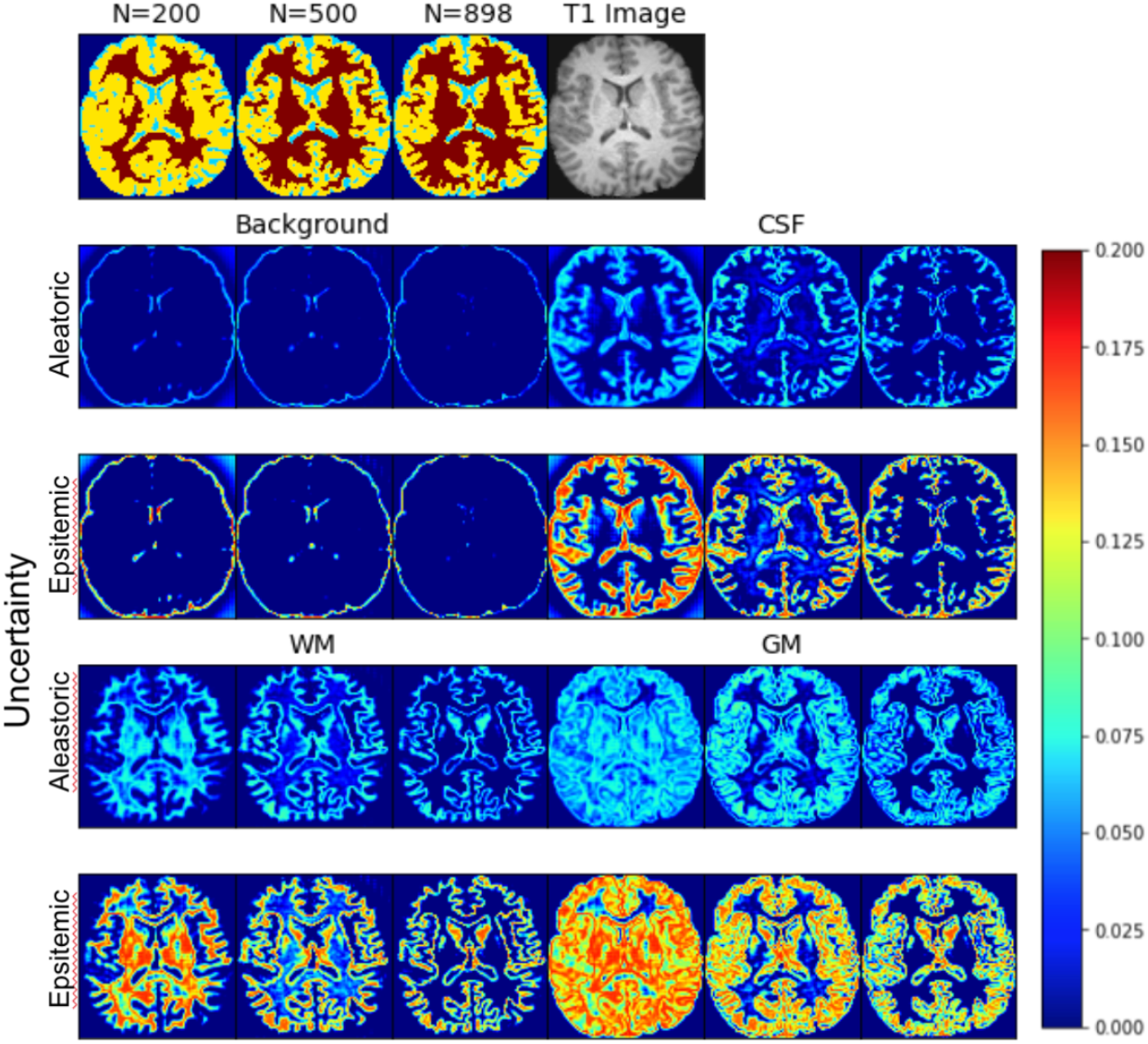
Tissue class prediction and MPRAGE image (top row). The aleatoric and epistemic uncertainty are shown for each tissue class. For each set of three the first is based on the N=200 model, the second on the N=500 model and the third based on the N=898 model. Data shown here from one slice from the connectome dataset (and was part of the test dataset).

Figure 3 shows the epistemic uncertainty (left) and aleatoric uncertainty (right), of the white matter, from the cerebellum to the top of the brain for the N=898 trained mode for a single test dataset. The highest epistemic uncertainty was primarily in the cerebellum. The aleatoric uncertainty was consistently lower than the epistemic uncertainty. There was a correspondence between increased epistemic uncertainty and increased aleatoric uncertainty.

**Figure 3:**
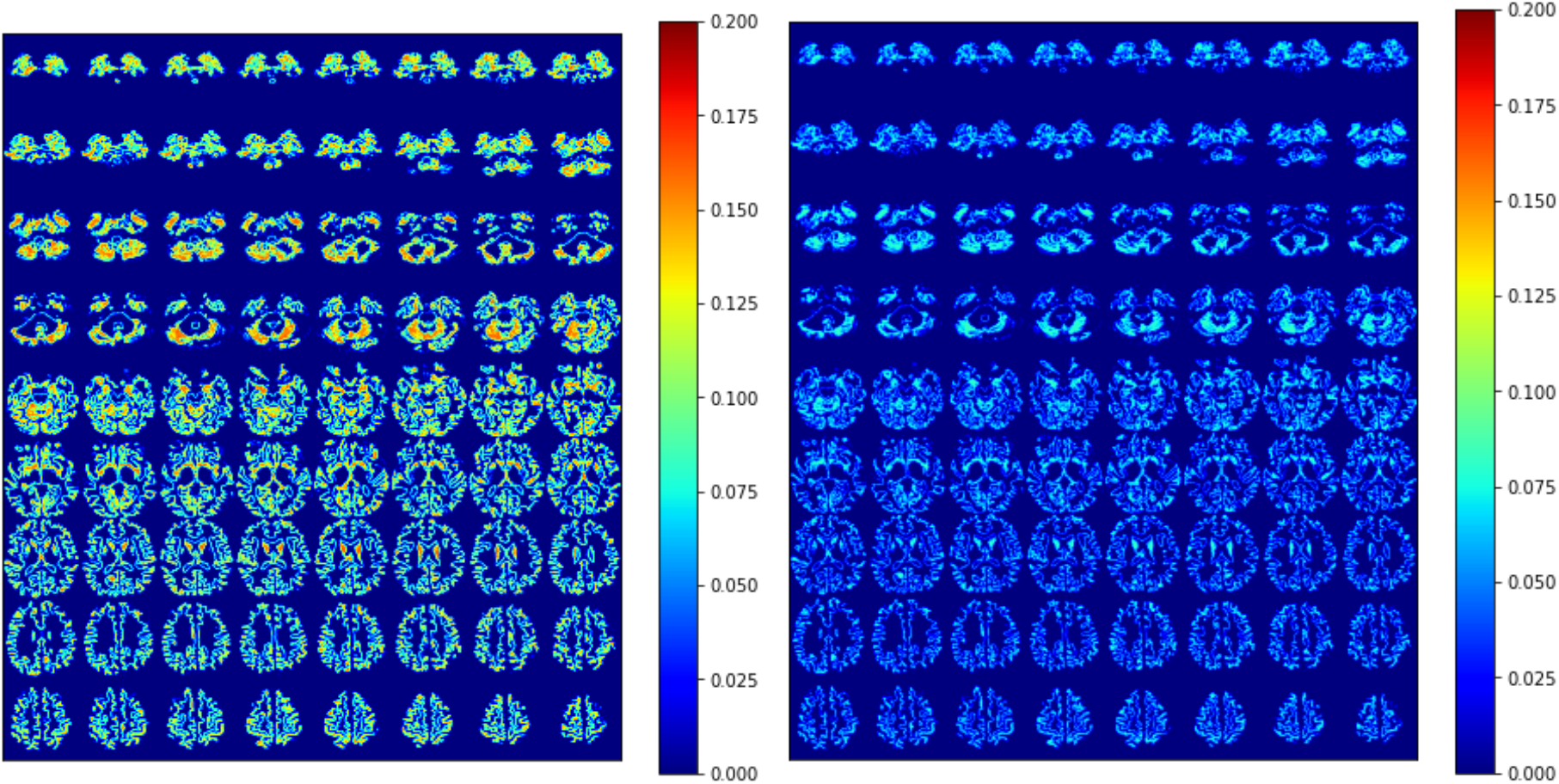
Whole brain epistemic uncertainty (left) and aleatoric uncertainty (right) of the white matter for one example volunteer scan from the 1000 connectome data.

### Brain Tumor Results

Figure 4 shows one slice through a glioma brain tumor along with the predicted tissue labels as well as the data and epistemic uncertainty maps. In general, there is a similar trend in the epistemic uncertainty and aleatoric uncertainty as was seen in a normal control (Figure 4). Much of the uncertainty is at the tissue boundaries (at least for the higher N). The aleatoric and epistemic uncertainty at N=200 had the highest uncertainty in most of the tissue types and in the region of the tumor. The tumor region had a signal appearance of CSF and was labeled as such by the network. The N=200 and N=500 epistemic and aleatoric uncertainty maps showed high uncertainty in the region of the tumor. The N=898 epistemic and aleatoric uncertainty maps, though, were much lower. This is likely due to the signal in the tumor being very similar to the CSF and therefore was predicted as such with low uncertainty.

**Figure 4:**
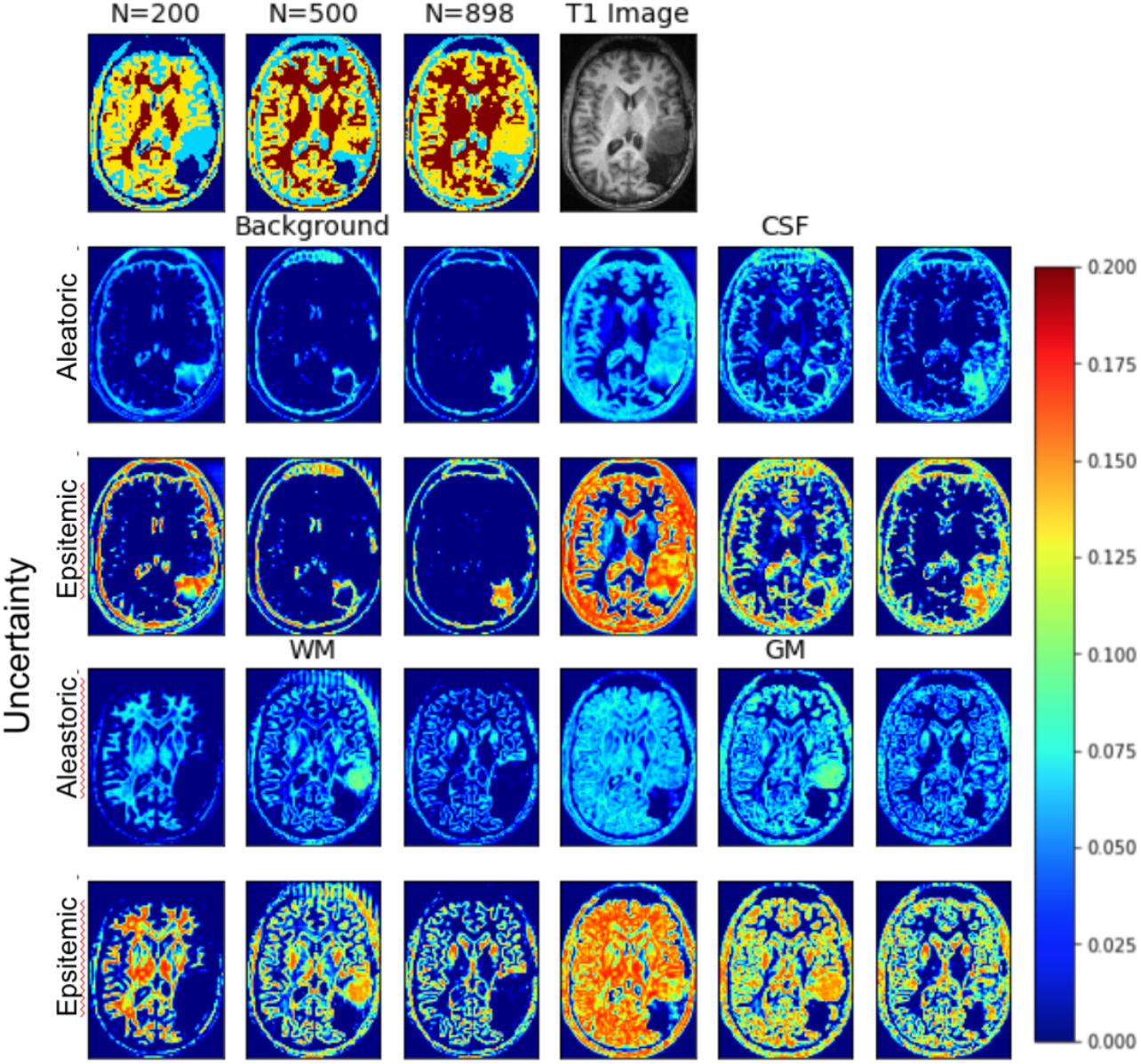
Tissue class prediction and MPRAGE image (top row). The data and epistemic uncertainty are shown for each tissue class. For each set of three the first is based on the N=200 model, the second on the N=500 model and the third based on the N=898 model. Data shown for one example tumor patient from a slice through the middle of the tumor.

### Aggregated Results

The aleatoric and epistemic uncertainty were averaged for each tissue over all test data as a function of the number of training sets for the model and is shown in Figure 5. The top panel shows the results for the test data and the bottom panel shows the results for the tumor data. Overall epistemic uncertainty was approximately twice that of the aleatoric uncertainty. The epistemic uncertainty also followed a decreasing trend as a function of the number of training sets. The aleatoric uncertainty had more variability and even though there appeared to be a decreasing trend it was small and likely just due to statistical fluctuations.

**Figure 5:**
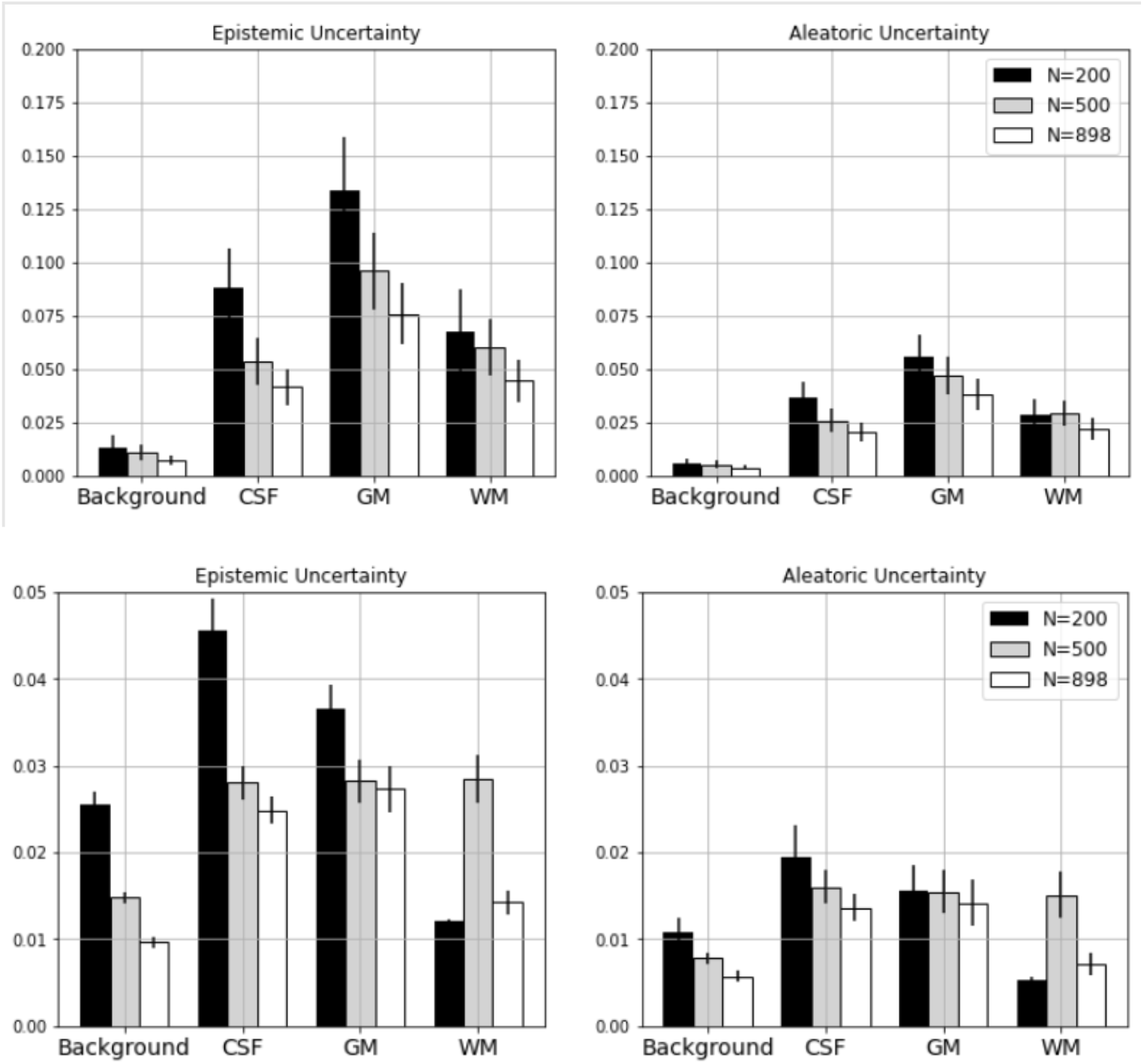
Mean epistemic and aleatoric uncertainty over all test images for each tissue class as a function of the trained model N=200 (black bars), N=500 (gray bars), N=898 (white bars). MPRAGE test images from 1,000 connectome project on top and tumor data on the bottom.

## Discussion

The goal of this study was to quantify the epistemic and aleatoric uncertainty directly from the results of the neural network segmentation without need of repeated measurements, bootstrap quantification, or other advanced techniques. There are numerous datasets out there that have a similar set of T1 MRI scans including Human Connectome Project, OpenNeuro.org, and OASIS. The 1000 Connectome dataset was chosen as it was a large and consistent set of MRI volumes. The neural network was trained with N=200, N=500 and N=898 T1 brain data from a public dataset and the accuracy and loss during training followed an expected trend. The epistemic and aleatoric uncertainty were quantified for each tissue class from each trained neural network in a test dataset and in a tumor image acquired at our site. The mean epistemic and aleatoric uncertainty were quantified for all control data and all tumor aleatoric and the epistemic uncertainty followed the expected trend of decreasing uncertainty as a function of number of data used to train the network.

This technique provides a simpler algorithm to quantify the data and epistemic uncertainty from a neural network prediction. There is not a requirement to assume prior distributions as required by some Bayesian inference models (Gelman et al. 2008). Nor is there a requirement to place priors on the network weights (Kingma, Salimans, and Welling 2015). The technique derived here is an extension to a derivation to estimate the epistemic and aleatoric uncertainty in a classification model (Amini et al. 2019) and we extended their work to a segmentation neural network. This extension should be appropriate for any type of U-Net as it only requires a change to the definition of the loss function.

The model loss was expected to decrease the more data added for training and the images showed this to be the case. It was expected the aleatoric uncertainty would be more consistent with increasing N but here the aleatoric uncertainty decreased as well. This can be explained by the consistency property of the maximum likelihood estimator. As the estimation of hyperparameters through minimizing the objective function can be seen as an analogy of maximum likelihood estimator (MLE), the consistency property applies here (Kiefer, et al. 1956). Moreover, the uncertainties are related to the hyperparameters through one-to-one functions (equation 4 and 5). The estimation of uncertainties converges to the truth in the order of 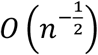. It corresponds to the results in Figure 5.

The main limitation to this technique, as implemented, is that it relies on signal comparison of a single signal intensity and therefore the uncertainty does not detect errors in segmentation class assignment. For example, the tumor segmentation in Figure 4 has a region with signal intensity similar to gray matter, and a region with a signal intensity similar to white matter. To minimize errors in this type of misclassification, a second complementary input could be added for more accurate classification. Alternatively, a spatial constraint may help to minimize the misclassification and therefore low uncertainty in regions mis-classified. The loss function derived and used in this study should be applicable for all segmentation networks and imaging types, but other networks were not tested. The gold standard relied on in this paper was based on a binary segmentation of the tissues in the brain and this may bias the uncertainty to be at the tissue boundaries more than necessary. A fuzzy segmentation may provide better segmentation results and lower uncertainty throughout the brain. There are options, though, to add other terms to the loss function which could add regularization or restrictions on the class definitions. The goal of this study however was to estimate epistemic and aleatoric uncertainty, not to optimize segmentations and in a sense, imperfect segmentations provide more information about the estimations.

Overall, the data and epistemic uncertainty were quantified per voxel throughout the whole brain based on the loss function implemented. The uncertainties were quantified directly from the U-Net output and did not rely on a bootstrap technique or assumptions about the underlying data. This uncertainty information could provide extra information for a radiological interpretation or may be used to provide an error measurement on the mask size. As the estimate of aleatoric uncertainty is statistically consistent, an estimate of this uncertainty would necessitate adequate sampling of data and its variance. A benefit of estimating the epistemic uncertainty is the assessment of whether increasing data samples or optimizing the neural network could potentially improve accuracy of the model. In situations where epistemic uncertainty is low, and yet the model does not reach intended targets for accuracy, it may be futile to further attempt to generate a model that may be of utility.

For medical applications, the accuracy of deep learning models needs to be evaluated (Senge et al. 2014) in order to determine the confidence level of the output for a particular model, whether it is for classification, segmentation, or other output. Specifically for segmentation, the inherent uncertainty of the segmentation output at the voxel level would potentially provide critical information to determine reliability of the data for medical decision making, for example in cases where accurate tumor volumes may be necessary to determine treatment.

## Conclusion

The U-Net segmentation neural network was able to be trained on the loss function derived and maps of data and epistemic uncertainty were created. The data and epistemic uncertainty followed a trend of decreasing uncertainty as the number of data used to train the neural network increased. The segmentation aleatoric uncertainty was primarily at the tissue boundary as the N increased, as expected. Overall, we believe this is a significant step forward for quantifying the uncertainty in neural network segmentations.

## Data Availability

The training and test data was used from the publicly available 1,000 Connectome project https://www.nitrc.org/projects/fcon_1000/. A small set of institutional tumor data was used for further algorithm testing but this data will not be available publicly.

## Appendix

### Loss Derivation

The multi-class classification problem with one-hot encoding notation and suppose we have *m* classes. The observation that a voxel belongs to the *j*th class is denoted a vector *Y*_*j*_with value 1at the *j*th element and 0 for all other elements. We assume *Y*_*j*_ ∼ *Multinomial*(*θ*), for *θ* = (*θ*_1,_, *θ*_2,_ …, *θ*_*m*_)^*T*^, whereclasses. The observation that a voxel belongs to the *j*th class is denoted a vector *Y*_*j*_ with value 1at the *j*th element and 0 for all other elements. We assume *Y*_*j*_ ∼ Multinomial(*θ*), for *θ* = (*θ*_1,_, *θ*_2,_ …, *θ*_*m*_)^*T*^, where

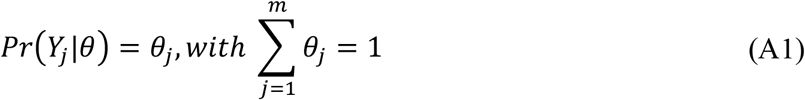

The Dirichlet distribution prior was assigned to *θ* is the Dirichlet distribution, with hyperparameters *α* = **(*α***_**1**_, ***α***_**2**_, **…**, ***α***_***m***_ )^***T***^and *α*_*j*_ > 0. The probability density function of the prior distribution is: is the Dirichlet distribution, with hyperparameters *α* = (α_**1**_, ***α***_**2**_, **…**, ***α***_***m***_ )^***T***^and *α*_*j*_ > 0. The probability density function of the prior distribution is:

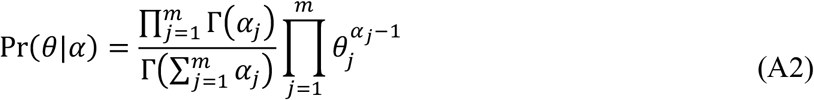

Given the distribution and probability density function the prediction is defined as

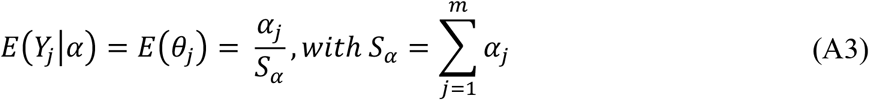

where the *α*_*j*_ are the tissue probability predictions used to quantify the tissue class maps. As well, the aleatoric uncertainty is derived as:

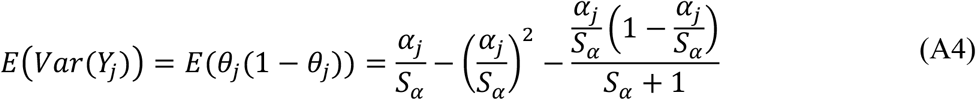

and the epistemic uncertainty is derived as:

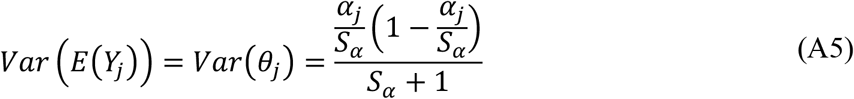

And the total uncertainty is *VarP*(*Y*_*j*_) = *Var* =(*E*(*Y*_*j*_|*θ*)) +*E* (*Var* (*Y*_*j*_|*θ*)).

The marginal distribution of Y given *α* can be derived by integrating out *θ* from the joint distribution, which has the following form,

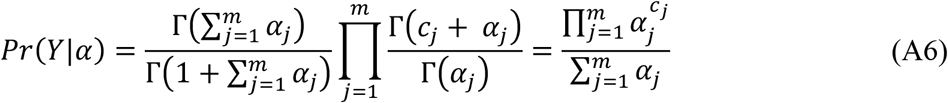

where *Г*(*c*) denotes the Gamma function, and *c*_*j*_ = 1 if *Y* belongs to the *j*th class, 0 otherwise. Then the loss function can be calculated as the negative logarithm of the likelihood defined by Equation A6:

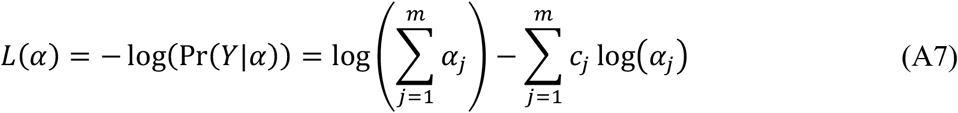

Therefore, the loss function defined in Equation A7 is the loss function used in the neural network training.

